# A counterfactual analysis quantifying the COVID-19 vaccination impact in Sweden

**DOI:** 10.1101/2024.08.22.24312361

**Authors:** Fanny Bergström, Felix Günther, Tom Britton

**Author notes:** Felix Günther changed affiliation after his work on this project and now works at the Department of Infectious Disease Epidemiology, Robert Koch Institute, Germany.

## Abstract

**Background:** Vaccination was the single most effective measure in mitigating the impact of the COVID-19 pandemic. Our study aims to quantify the impact of vaccination programmes during this initial year of vaccination by estimating the number of case fatalities avoided, having Sweden as a case study.

**Methods:** Using Swedish data on age-specific reported incidence, vaccination uptake, and contact structures, along with age-specific estimates on the vaccine efficacies and under-reporting from the literature, we fit a Bayesian SEIR epidemic model with time-varying community contact rate *β*(*t*) for COVID-19 incidence. By adding age-specific infection fatality rates, we obtain an estimate of about 5,540 (95% PI: 5,390-5,690) for the number of case fatalities from the fitted model. This estimate aligns closely with the reported 5,430 case fatalities during the same period. We then use the estimated contact rate *β*(*t*) in a counterfactual analysis where the population is unvaccinated, leading to more infections and fatalities.

**Findings:** The counterfactual analysis result in a severe epidemic outbreak during the early autumn of 2021, resulting in about 52,600 (49,900-55,500) number of case fatalities. Consequently, the number of lives saved by the vaccination program is estimated to be about 47,100 (44,500-49,800), out of which 6,460 are directly saved and 40,600 are indirectly saved, mainly by drastically reducing the severe outbreak in the early autumn of 2021, which would have occurred without vaccination and unchanged community contact rate.

**Interpretation:** Our mathematical model is used to analyze the impact of COVID-19 vaccination on lives saved in Sweden during 2021, but the same methodology can be applied to other countries. The counterfactual analysis offers insights into an alternative trajectory of the pandemic without vaccination. By incorporating estimated vaccination-related parameters, age-specific infection fatality ratio and under-reporting, our model estimates the number of case fatalities avoided. The results not only show the direct impact of vaccination on reducing deaths for infected individuals but also shed light on the indirect effects of reduced transmission dynamics.

**Funding:** NordForsk (project number 105572).

**Research in context:** *Evidence before this study:* Vaccination against COVID-19 has been proven to reduce infection rates, hospitalizations, and mortality. Observational studies and clinical trials have demonstrated the efficacy COVID-19 vaccines, and mathematical modeling studies have been conducted to predict the potential outcomes of vaccination programmes under different scenarios. These models have provided insights into the benefits of achieving high vaccination coverage and the consequences of delays or interruptions in vaccine distribution. There remains a gap in analyses that quantify the impact of the vaccination programme in Sweden by comparing actual outcomes with a scenario without vaccination. This study aims to fill this gap by employing a robust counterfactual analysis method to provide a clearer picture of the COVID-19 vaccination impact in Sweden. This approach offers a understanding of the impact of vaccination by isolating the effects of the vaccination from other interventions. For our literature review, we have searched PubMed for “vaccination counterfactual analysis”, “SEIR model vaccination impact”, “quantifying effect of COVID-19 vaccination” and “COVID-19 IFR”. The Swedish Public Health Agency has provided country-specific data for Sweden.

*Added value of this study:* The Bayesian SEIR model presented in this paper provides a flexible and data-driven framework to assess the effectiveness of vaccination strategies. Using age- and country-specific data and parameters on reported cases, under-reporting and infection fatality rate (IFR), we can quantify the effect of vaccination in Sweden 2021. This study compares the case fatalities in a factual analysis with a counterfactual analysis, which has the same contact rates but with a unvaccinated population. The comparison allows an estimate of the number of lives saved from the vaccination. The methodology can be used to evaluate the effect of vaccination in other countries, but also for other counterfactual scenarios such as different vaccination schemes.

*Implications of all the available evidence:* Our findings highlight the critical role of vaccination in mitigating COVID-19-related mortality. Through the counterfactual analysis provided by the SEIR epidemic model, we gain insights into the effects of vaccination programmes. Beyond reducing deaths directly attributable to COVID-19 infection vaccination also exerts a broader societal impact by curbing transmission rates and easing strain on healthcare systems. Moreover, by quantifying the number of lives saved through vaccination efforts, we offer policymakers and public health officials invaluable data for optimizing future vaccination strategies and reinforcing the importance of widespread vaccine uptake. The insights gained from this study not only show the effectiveness of vaccination in saving lives but also provide a robust framework of a data-driven approach to guide evidence-based decision-making and shaping vaccination policies.

## Introduction

COVID-19 vaccination campaigns are credited with saving an estimated 20 million lives worldwide in their first year of deployment only.^1^ This life-saving impact is twofold: directly, by reducing severe illness and preventing deaths upon infection, and indirectly, by lowering transmission rates among vaccinated individuals.^2^ Watson and colleagues ^1^ conducted a comprehensive study assessing the global impact of the initial year of vaccination where they estimate the direct and indirect effects but do not take into account the age-specific transmission rates and infection fatality ratio (IFR). Also, country-specific efforts have been made to quantify the impact of vaccination directly^3*−*5^ and both directly and indirectly.^6,7^

Mathematical modelling is a tool that allows simulation of possible alternative trajectories of the pandemic through counterfactual analyses.^8,9^ In this paper, we are interested in quantifying the effects of vaccination on COVID-19 fatalities in Sweden during 2021. The reason for focusing on 2021 is that vaccination started close to January 1st 2021, and in late December of 2021 Omicron started to take off with less vaccine efficacy as well as severity, complicating the analysis. We investigate the effect of vaccination by performing a retrospective analysis of COVID-19 transmission dynamics and comparing estimated dynamics from observed disease burden to a counterfactual simulation on alternative transmission dynamics and expected disease burden in the hypothetical scenario without vaccination. Utilizing data on COVID-19 incidence, vaccination coverage, and demographic variables, the Susceptible-Exposed-Infectious-Removed (SEIR) epidemic model can capture the dynamic interplay among individuals in the population. This offers insights into what could have happened in the alternative scenario. We keep other restrictive measures and interventions constant when evaluating the effect of vaccination to isolate its effect.

This paper provides a framework for quantifying lives saved through COVID-19 vaccination efforts in Sweden. We estimate the effect of direct protection among vaccinated individuals and the indirect effect due to reduced transmission in society. Specifically, we examine trajectories of COVID-19 in scenarios with and without vaccination while holding contact patterns constant.

## Methods

### Analysis

A time-varying contact intensity *β*(*t*) is estimated from reported cases using an age-structured Susceptible-Exposed-Infectious-Removed (SEIR) compartmental model including a social contact matrix and time varying and age-specific vaccination uptake as well as externally estimated under-reporting and vaccine efficacies acting on reducing susceptibility, infectivity and mortality.

time varying and age-specific vaccination uptake as well as externally estimated under-reporting and vaccine efficacies. In a counterfactual analysis, we use the *β*(*t*) estimated in the factual analysis previously described. Using this *β*(*t*) we simulate an age-structured SEIR epidemic model with the same social contact matrix but in a population where no one is vaccinated. The difference between the simulated disease burden from the counterfactual analysis and the factual analysis can be interpreted as the effect of the vaccination programme since we assume everything equal except for the vaccination in the two simulations.

In an intermediate model we use the same infection pressure as estimated in the main analysis, i.e. assuming the same *β*(*t*) and the same numbers of infectives of different age groups as well as vaccinated and unvaccinated individuals. Given this infection pressure over time, we simulate how many would get infected in the different age groups over time, again assuming that no one was vaccinated but with the difference that this infection pressure is decoupled from how many get infected in the model (so infected individuals do not contribute to new infections) and the infection pressure can better be viewed as an external infection pressure. As a result, the infection pressure in the intermediate analysis remains as low as in the factual analysis. This allows us to interpret the difference in the simulated number of case fatalities from the intermediate and factual analysis as the direct effect of the vaccination programme. We derive the remaining effect of vaccination from the difference in the counterfactual and intermediate analysis, which we interpret as the indirect effect.

The SEIR model provides estimates of the number of infections in the different age groups in the three scenarios; factual, counterfactual and intermediate. We use these estimates of the number of infections together with the age-specific IFR and vaccine efficacy against mortality to get the estimated case fatalities. We thereby use the difference between these estimates from the factual, intermediate and counterfactual analysis to quantify the effect of vaccination. We use a Bayesian framework to account for uncertainties in parameter estimates, providing a robust foundation for assessment.

### SEIR model

We consider a constant population size, at any time *t*, of *N* individuals but allow for imported cases. We divide the population into age groups of 10-year spans; *a* = 0 *−* 9, 10 *−* 19, …. The population is further divided into four compartments or classes. These are called the susceptible, exposed, infectious and removed classes. Their sizes, at any time *t* for an age class *a*, are denoted by *S*^(*a*)^(*t*), *E*^(*a*)^(*t*), *I*^(*a*)^(*t*) and *R*^(*a*)^(*t*) respectively. We let the rate at which individuals become infectious after being latently infected be denoted *θ* and the recovery rate from the infectious state *γ*. We use a contact matrix *C* estimated by Prem et al.^10^, with elements *C*_*a′,a*_ being the average number of contacts an individual in age group *a*^*′*^ has with individuals in age group *a* within 24 hours. We also use the time-varying overall community contact rate *β*(*t*). We specify the full SEIR model in the Supplementary Material, Section 1.

Subscript *u, v*1 and *v*2 are used to distinguish between the unvaccinated and vaccinated groups with one or two doses. We further let *ve*_*susc*1_, *ve*_*susc*2_, *ve*_*inf*_ and *ve*_*mort*_ be the vaccine efficacy in terms of susceptibility with one and two doses, infectiousness and mortality. We consider a leaky vaccine,^11^ meaning that susceptible vaccinated individuals have a reduced probability of being infected upon an infectious contact by (1 *− ve*_*susc*1_) with one dose of vaccination and (1 *− ve*_*susc*2_) with two doses of vaccination. We assume a reduced rate of infecting others by (1 *− ve*_*inf*_) regardless of the number of vaccination doses if infected. The reduced risk of dying if infected is 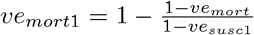 and 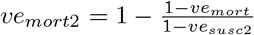 for one and two doses respectively. The number of deaths at time *t* in age group *a* is calculated from the newly exposed individuals *n*(*t*) as

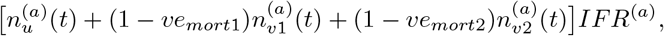

without time delay from time of infection to time of death.

In the factual model, the infection pressure at time *t* for an unvaccinated individual in age group *a*, with a population size of *N*_*a*_, is defined as

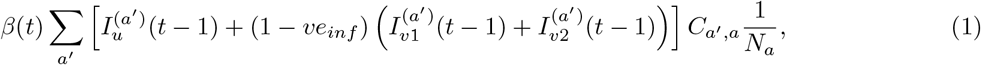

where *β*(*t*) is estimated within the model, and infectious individuals contribute to new infections. In the counterfactual model, where everyone remains unvaccinated, the infection pressure for an individual in age group *a* is defined

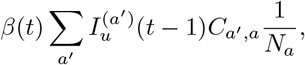

with *β*(*t*) from the factual model. In the intermediate model, the infection pressure is identical to the factual analysis (i.e. Eq (1) evaluated in the factual analysis), but all uninfected individuals are fully susceptible.

### Data sources

In 2021 there were 882,000 cases, 4,100 admissions to the ICU and 5,530 case fatalities (including all age groups) reported in Sweden.^12^ During 2021 the Sars-Cov-2 variants Ahlpa (February-June) and Delta (July-December) mainly circulated in Sweden, but in early January 2021, it was still Wildtype and the end of December 2021 was the start of the Omicron wave.^15^

The amount of under-reporting of cases depends on the behavior of the population and testing strategy which varied over time. To account for this we use estimated under-reporting from the Public Health Agency (PHA) of Sweden.^16^ The PHA provided two estimates; model-free and model-based (see Supplementary Material, Section 2.2). We use the model-free estimates of the under-reporting in our main analysis and the alternative model-based estimates in a sensitivity analysis. We assume that the initial number of immune is the cumulative number of reported cases before 2020–12–31 up-scaled by the age-specific under-reporting, resulting in about 11% of the population being in the R (recovered) compartment at the start of 2021.

We use the time series of infections (up-scaling reported incidence using the under-reporting estimates) for the analysis, seen in Figure 1A. This quantity is derived from the reported cases divided by the estimated fraction reported for each age group. We use the day of reporting as the day of getting infected, which in reality occurs before. As previously described we also disregard the delay from getting infected until time of death, which occur later. In other words, we disregard two delays in opposite directions. For the time series of reported cases, we use a 7-day rolling average to circumvent the weekday effect of reporting. We exclude age group 0-19 from our analysis, as testing in this age group was very limited in Sweden.

**Figure 1:**
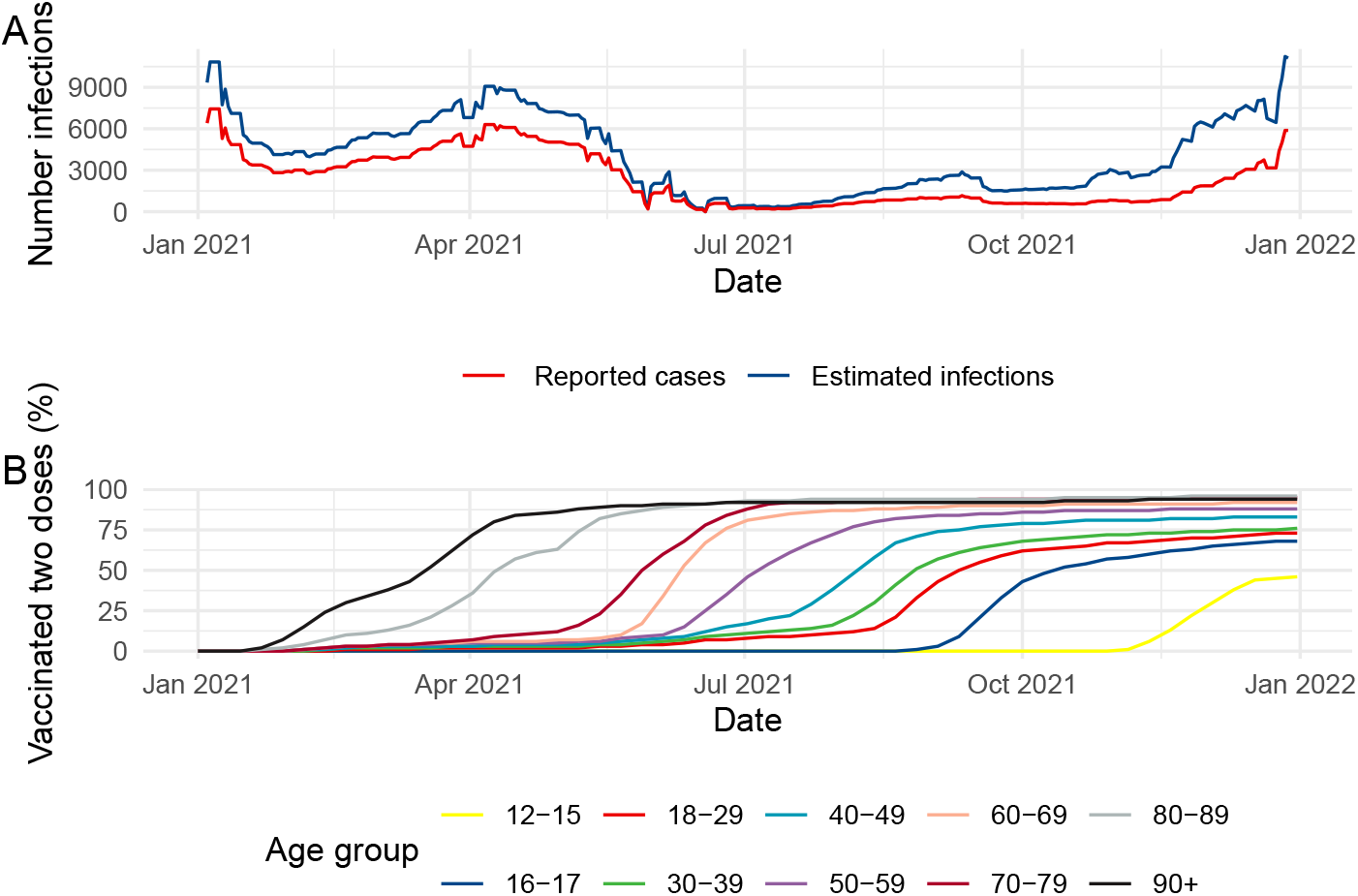
Reported cases (red) and estimated number of infections (blue) derived from the reported cases up-scaled by the under-reporting with no assumed delay from time of infection to time of reporting (A). Fraction of COVID-19 vaccinated individuals in Sweden during 2021 per age group as per date of receiving the second vaccination dose (B).

The COVID-19 vaccination started in Sweden 2020–12–27. It was distributed so that people in risk groups and 80+ years of age had priority and later opened up for younger age groups in descending order for 10-year age groups, 70-79 followed by 60-69, 50-59 until the youngest age groups (Figure 1B).

We use age-specific IFR estimates derived from the literature.^17*−*19^ We use the combined information from the three sources as no source we have found provides estimates for each age group used in our analysis. Data on the demography of Sweden is collected from Statistics Sweden.^20^

The vaccine effectiveness (VE) values against mortality used in the analysis is 91%, accounting for reduced risk of infection and dying if infected. This is the same used for two doses of vaccination as used by the WHO European Respiratory Surveillance Network.^21^. We also use the same effectiveness regardless of the vaccination type and variant (during 2021). The VE in our analysis in terms of infectiousness is 50% and for susceptibility 50% and 80% for one and two doses of vaccination respectively.^22^ The mean of duration of incubation period (*θ*^*−*1^) 4.6 days and mean duration of infection 2.1 (*γ*^*−*1^) is the same values as in Hogan et al.,^23^ resulting in a mean generation time of 6-7 days. The age-specific IFR and other parameters used in our analysis are found in Supplementary Material, Section 2.

### Statistical analysis

Programming language R version 4.2.1^24^ is used for the analysis. The Bayesian hierarchical model is implemented in Stan^25^ and used with CmdStanR.^26^ The code and data used for our analysis are publicly available at https://github.com/fannybergstrom/vaccination_sweden. Further details on the statistical analysis and the Bayesian model are found in Supplementary Material, Section 3.

## Results

The factual analysis results in about 949,000 (95% PI: 921,000-978,000) number of infections in 2021, meaning that 12% of the population were infected during this time. The estimated number of case fatalities in the factual analysis is 5,540 (95% PI: 5,390-5,690), which aligns closely with the official fatality count of 5,430 reported for individuals over 20 years old in Sweden during the same period. Figure 2A illustrates the reported and estimated case fatalities from the factual analysis over time, from we can see that our model captures the trend of case fatalities but with a slight underestimation in the first months of 2021 and an overestimation from March to May. This discrepancy may be due to variations in under-reporting or changes in the infection fatality rate (assumed to be constant over time). The analysis is stratified by 10-year age groups, but the results presented in Figures 3 and 4 are aggregated. Detailed results for all age groups are available in Table 1 and Supplementary Material, Section 4.

**Table 1:**
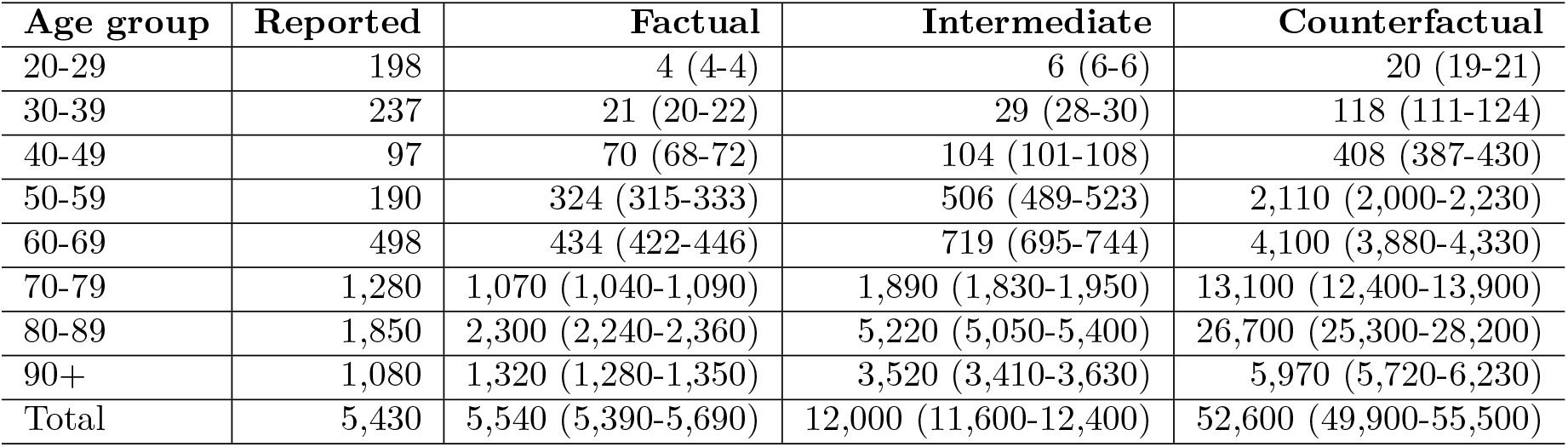
Reported and estimated number of case fatalities per age group from the factual, intermediate and counterfactual analyses. Shown are the median of the posterior predictive distribution and the 95% PI.

**Figure 2:**
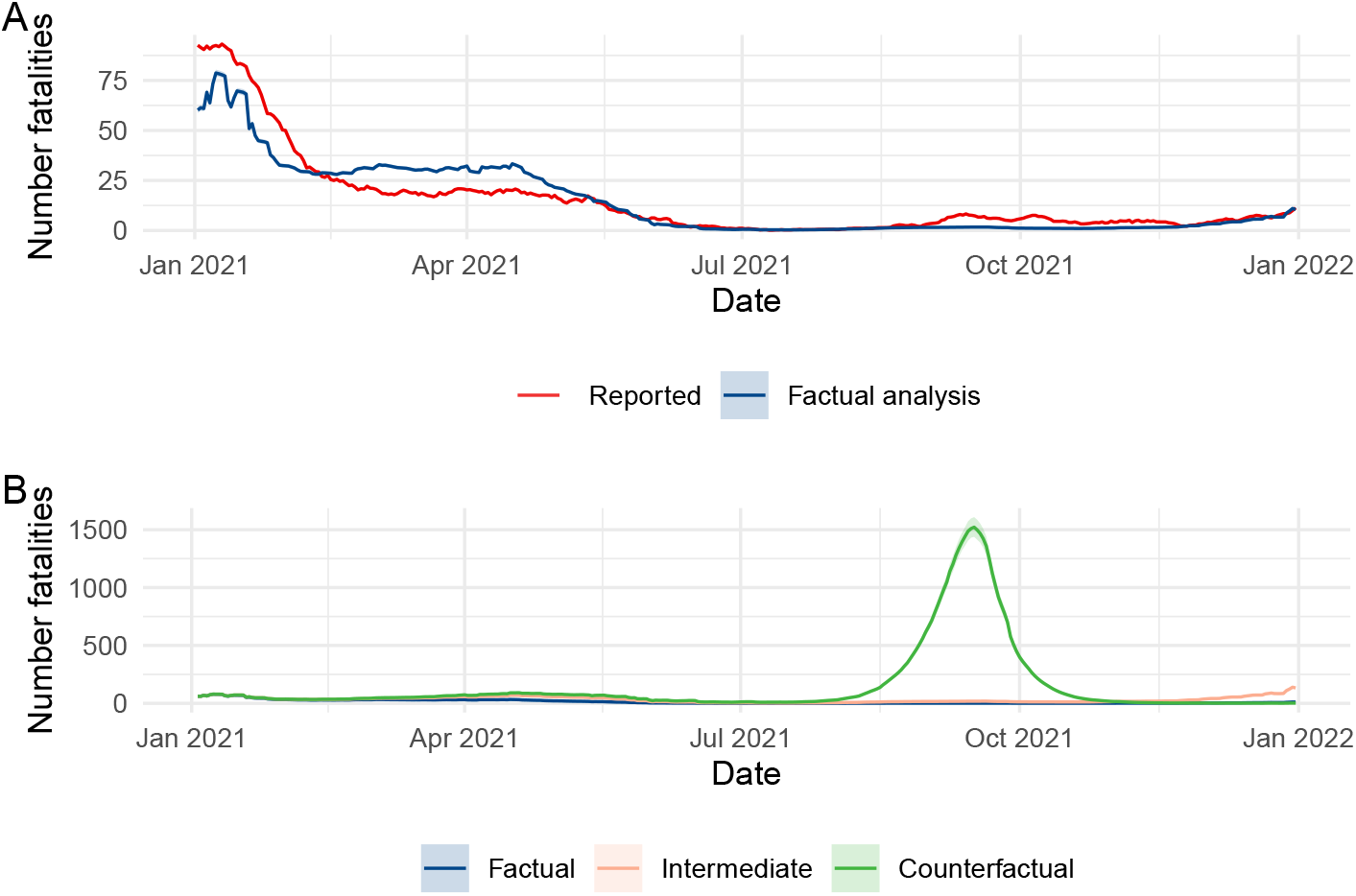
Reported and estimated fatalities from factual, intermediate and counterfactual models. The number of estimated case fatalities in the factual analysis can be seen to agree fairly well with the official fatality count (A). The counterfactual analysis result in a severe outbreak in early autumn (B). Please note the very different scales on the Y-axis of the two sub-figures. The blue curve in the two subfigures are the same.

In the counterfactual analysis, we assume that no one was vaccinated and that the community contact rate *β*(*t*) remained the same as when vaccination occurred. This results in a severe epidemic outbreak during the early autumn of 2021. This would have led to only 20% of the population escaping infection by the end of 2021, leading to herd immunity (in the absence of immunity waning and an influx of new susceptible individuals). The counterfactual model results in about 6,170,000 (5,850,000-6,500,000) estimated number of infections, an increase of 550% compared to the factual analysis, leading to about 52,600 (49,900-55,500) number of case fatalities. In the intermediate analysis we estimate about 12,000 (11,600-12,400) number of case fatalities. In other words, we estimate that about 47,100 (44,500-49,800) number of lives were saved in Sweden during 2021, combining the direct and indirect effects of vaccination.

In late 2021 there is a rise in number of case fatalities in the intermediate model while in the factual and counterfactual model it is remained low, seen in Figure 2B. This implies that the infection pressure is rising but since in the factual model the majority of the population is protected through vaccination and in the counterfactual model from natural immunity from infection, therefore the case fatalities remain low in these two models. Without a broad vaccination programme and a COVID wave in autumn 2021 that was kept relatively under control (e.g., through substantial contact reduction), a large proportion of the population would still have been susceptible to infection. Hence there would still be high risk for a large outbreak and a significant associated disease burden in the end of 2021 and the following months.

The total estimated number of case fatalities agrees with the official fatality counts, as shown in Figure 2A. These estimates also align reasonably well with the reported deaths stratified by age seen in Table 1. The table further reveals that we, in the factual analysis, underestimate the deaths in the younger population and overestimate the deaths among the older but to a smaller extent. Overall in the age groups over 40 years, our estimates in the factual analysis aligns well with the reported numbers.

We measure the indirect effect as the difference between the counterfactual and intermediate analysis and the direct effect as the difference between the intermediate and the factual. The number of lives saved by the vaccination program is hence estimated to about 47,100 (95% PI: 44,500-49,800), out of which 6,460 are directly saved through protection among vaccinated individuals by reducing their risk of infection and mortality and 40,600 are indirectly saved by the reduced infection pressure. At the beginning of the year when it is mainly the older population that is vaccinated, the direct effect is relatively larger than for the second part, where it is the indirect effect driving the effect of vaccination. The estimated effect of vaccination divided into the first and second half of 2021 is found in Table 2. We chose this time point since in the first half of the year it is mainly the population of age 60+ that was vaccinated (seen in Figure 1B) which is also the age groups with significantly higher IFR.

**Table 2:** Estimated number of case fatalities avoided from vaccination in terms of direct, indirect and total effect. The results are further separated into two time periods; before and after 2021–07–01.

| Time | Direct | Indirect | Total |
| --- | --- | --- | --- |
| All year | 6,460 (6,210-6,710) | 40,600 (38,300-43,100) | 47,100 (44,500-49,800) |
| Before 2021-07-01 | 2,630 (2,550-2,700) | 1,790 (1,740-1,850) | 4,420 (4,290-4,550) |
| From 2021-07-01 | 3,830 (3,660-4,010) | 38,800 (36,500-41,200) | 42,600 (40,200-45,200) |

We estimated case fatalities based on the number of infections, without considering a delay between the time of infection and death. We also disregarded the delay from the time of infection to reporting. If we had assumed a 5-day delay from infection to reporting and a 12-day delay from infection to death, we would have shifted the number of reported case fatalities by one week. This adjustment would result in an estimated 5,020 reported case fatalities for one year, compared to the reported number 5,430. This number also aligns reasonably well with our factual analysis.

### Sensitivity analysis

As a sensitivity analysis, we repeat the main analysis but vary the assumptions made. The relative impact in terms of number of lives saved by varying one parameter while keeping the remaining as in the main analysis is found in Table 3. Changing the mean duration of the incubation period (*θ*^*−*1^) and infectious period (*γ*^*−*1^), and the number of imported cases has little impact on the results. However, changing the assumptions about the under-reporting, number of initial immune and IFR has a more significant impact on the analysis.

**Table 3:** Results from sensitivity analysis. Percentage change in the number of case fatalities compared to the main analysis when varying one parameter.

| Parameter | Change (%) | Direct (%) | Indirect (%) | Total (%) |
| --- | --- | --- | --- | --- |
| Duration of incubation period | -25% | 0.74 | -0.45 | -0.29 |
| Duration of incubation period | +25% | 0.76 | 0.07 | 0.17 |
| Duration of infection | -25% | 0.67 | -1.21 | -0.95 |
| Duration of infection | +25% | 0.84 | 0.78 | 0.78 |
| IFR | -25% | -25.0 | -25.0 | -25.0 |
| IFR | +25% | 25.0 | 25.0 | 25.0 |
| Imported cases | -25% | 0.48 | 0.10 | 0.16 |
| Imported cases | +25% | 0.98 | -0.44 | -0.24 |
| Initial immune | -25% | 13.5 | 4.28 | 5.54 |
| Initial immune | +25% | -10.2 | -5.45 | -6.10 |
| Underreporting | Model-based | 160 | -37.3 | -10.3 |

For the under-reporting, we use the model-free estimates in the main analysis and the model-based estimate in the sensitivity analysis. Using the model-based estimates of the under-reporting shifts a considerable weight of the number of lives saved to the direct effect. This is mainly driven by a larger fraction of unreported cases in the elderly population at the beginning of 2021 (in detail shown in Supplementary Material, Section 5).

## Data Availability

All data used for our analysis are available online at https://github.com/fannybergstrom/vaccination_sweden/

https://www.folkhalsomyndigheten.se/smittskydd-beredskap/utbrott/aktuella-utbrott/covid-19/statistik-och-analyser/

https://www.folkhalsomyndigheten.se/contentassets/da0321b738ee4f0686d758e069e18caa/skattning-letalitet-covid-19-stockholms-lan.pdf/

https://www.scb.se/hitta-statistik/sverige-i-siffror/manniskorna-i-sverige/befolkningspyramid-for-sverige/

## Discussion

We use a mathematical model to analyze the impact of COVID-19 vaccination on lives saved in Sweden during 2021. Utilizing data on COVID-19 incidence, vaccination coverage, and demographic variables, the SEIR epidemic model captures the dynamic interplay among susceptible, exposed, infectious, and removed individuals, offering insights into the alternative trajectory of the pandemic in the case when there would have been no vaccination. A Bayesian framework is applied to account for uncertainties in parameter estimates, providing a robust foundation for assessment. This method can be used to evaluate the effect of vaccination in other countries and for other counterfactual scenarios, such as different vaccination schemes.

By considering country- and age-specific under-reporting, time of vaccination, IFR, and contact behaviour, our model facilitates the estimation of lives saved by COVID-19 vaccines in Sweden. The results not only emphasize the direct impact of vaccination on reducing the number of case fatalities but also the indirect effects on transmission dynamics. Our results indicate that in the early phase of vaccination, when the older population is mainly vaccinated, the direct effect is relatively more prominent than in the later stage when the indirect effect is the most prominent. We estimate that about 47,100 (44,500-49,800) number of lives were saved by vaccination, an increase of 13% compared to Watson et al.^1^

Our model takes several heterogeneities into account, in particular with respect to age. Still the model is a simplification of the actual spread of COVID-19, for example by not considering additional household transmission and other social structures, and also for modelling Sweden as one homogeneous community. One step toward realism would be to model spreading on a regional level, but there is a clear trade-off with doing this by also including more uncertainty in the model. Another potential problem is the various uncertainties in our parameter choices and even with our sensitivity analysis, a more extensive such could shed additional light on which assumptions are most influential. We do not account for waning of immunity from infection or vaccination. The time frame of our analysis is only one year but for longer simulations this would be an important aspect to include. A more specific consideration is that we estimate a time-varying community contact rate *β*(*t*) thought to reflect changes in behavior due to individual preventions as well as national recommendations. Some of these preventions, such as self-isolation and contact tracing, are also known to change the generation time distribution,^27,28^ and in the current modelling the latent and infectious period are assumed to remain constant in our analysis. To estimate changes over time for these quantities is of course very hard, and we believe such changes would not dramatically alter our conclusion. In reality, it is likely that additional behavior or non-pharmaceutical interventions such as restricted contacts would have reduced the magnitude of the severe outbreak occurring in our counterfactual analysis, but our comparison assumes everything but vaccination stays constant.

The Bayesian SEIR model presented in this paper provides a flexible and data-driven framework to assess the effectiveness of vaccination strategies (to be used also on other countries). This paper contributes to the literature focused on quantifying the impact of COVID-19 vaccination.

## Acknowledgement

We want to thank Paolo Scalia Tomba for helpful conversations and Sharon Kuhlmann with colleagues at the public health agency of Sweden for providing data. T.B. and F.G. acknowledge financial support from NordForsk (project number 105572).

