## Supplementaty_materials: Appendix for "A counterfactual analysis quantifying the COVID-19 vaccination impact in Sweden"

### Supplementary Materials: A counterfactual analysis quantifying COVID-19 vaccination impact in Sweden

Fanny Bergström[1], Felix Günther[1*], Tom Britton[1]
[1] Stockholm University, Stockholm, Sweden.

2024-08-15

#### 1 SEIR model

We consider a constant population size, at any time $t$, of $N$ individuals. We divide the population into age groups of 10-year spans; $a = 0-9, 10-19, \ldots$. The population is further divided into four compartments or classes. These are called the susceptible, exposed, infectious and removed classes. Their sizes, at any time $t$ for an age class $a$, are denoted by $S^{(a)}(t)$, $E^{(a)}(t)$, $I^{(a)}(t)$ and $R^{(a)}(t)$ respectively. We let the rate at which individuals become infectious after being latently infected be denoted $\theta$ and the recovery rate from the infectious state $\gamma$. We use a contact matrix $C$with elements $C_{a',a}$ being the average number of contacts an individual in age group $a'$ has with individuals in age group $a$ within 24 hours. We also use the time-varying overall community contact rate $\beta(t)$.

We let $\mathrm{n}_{vacc}(t)$ be the number of people with a new vaccination status of day $t$. Subscript $u$, $v1$ and $v2$ are used to distinguish between the unvaccinated and vaccinated groups with one or two doses. We further let $ve_{susc1}$, $ve_{susc2}$, $ve_{inf}$ and $ve_{mort}$ be the vaccine efficacy in terms of susceptibility with one and two doses, infectiousness and mortality. Susceptible vaccinated individuals have a reduced probability of being infected upon an infectious contact by $(1 - ve_{susc1})$ with one dose of vaccination and $(1 - ve_{susc2})$ with two doses of vaccination. We assume a reduced rate of infecting others by $(1 - ve_{inf})$ regardless of the number of vaccination doses if infected. We allow for imported cases and let $n_{imp}(t)$ is the number of imported cases of day $t$. We assume that the same number of imported infections and are removed from the susceptible compartment to keep the population size constant.

We specify the SEIR model as follows

$$\begin{aligned}S_u^{(a)}(t) &= S_u^{(a)}(t-1) - \mathrm{n}_{vacc1}^{(a)}(t-1)\frac{S_u^{(a)}(t-1)}{S_u^{(a)}(t-1) + R_u^{(a)}(t-1)} \\ &\quad - \beta(t)\sum_{a'} C_{a',a}\big(I_u^{(a')}(t-1) + (1-ve_{inf})(I_{v1}^{(a')}(t-1) + I_{v2}^{(a')}(t-1)\big)\frac{S_u^{(a)}(t-1)}{N_a} - n_{imp,u}^{(a)}(t)\end{aligned}$$

$$\begin{aligned}S_{v1}^{(a)}(t) &= S_{v1}^{(a)}(t-1) + n_{vacc1}^{(a)}(t-1)\frac{S_u^{(a)}(t-1)}{S_u^{(a)}(t-1) + R_u^{(a)}(t-1)} - n_{vacc2}^{(a)}(t-1)\frac{S_{v1}^{(a)}(t-1)}{S_{v1}^{(a)}(t-1) + R_{v1}^{(a)}(t-1)} \\ &\quad - \beta(t)\sum_{a'} C_{a',a}\big(I_u^{(a')}(t-1) + (1-ve_{inf})(I_{v1}^{(a')}(t-1) + I_{v2}^{(a')}(t-1)\big)\frac{S_{v1}^{(a)}(t-1)}{N_a}(1-ve_{susc1}) - n_{imp,v1}^{(a)}(t)\end{aligned}$$

[*]Felix Günther changed affiliation after his work on this project and now works at the Department of Infectious Disease Epidemiology, Robert Koch Institute, Germany.

$$
\begin{aligned}
S_{v2}^{(a)}(t) = S_{v2}^{(a)}(t-1) + n_{vacc2}^{(a)}(t-1)\frac{S_{v1}^{(a)}(t-1)}{S_{v1}^{(a)}(t-1)+R_{v1}^{(a)}(t-1)} \\
-\beta(t)\sum_{a'} C_{a',a}\big(I_u^{(a')}(t-1)+(1-ve_{inf})(I_{v1}^{(a')}(t-1)+I_{v2}^{(a')}(t-1)\big)\frac{S_{v2}^{(a)}(t-1)}{N_a}(1-ve_{susc2})-n_{imp,v2}^{(a)}(t)
\end{aligned}
$$

$$
E_u^{(a)}(t) = E_u^{(a)}(t-1)+\beta(t)\sum_{a'} C_{a',a}\big(I_u^{(a')}(t-1)+(1-ve_{inf})(I_{v1}^{(a')}(t-1)+I_{v2}^{(a')}(t-1)\big)\frac{S_u^{(a)}(t-1)}{N_a}-\theta E_u^{(a)}(t-1)
$$

$$
E_{v1}^{(a)}(t) = E_{v1}^{(a)}(t-1)+\beta(t)\sum_{a'} C_{a',a}\big(I_u^{(a')}(t-1)+(1-ve_{inf})(I_{v1}^{(a')}(t-1)+I_{v2}^{(a')}(t-1)\big)\frac{S_{v1}^{(a)}(t-1)}{N_a}-\theta E_{v1}^{(a)}(t-1)
$$

$$
E_{v2}^{(a)}(t) = E_{v2}^{(a)}(t-1)+\beta(t)\sum_{a'} C_{a',a}\big(I_u^{(a')}(t-1)+(1-ve_{inf})(I_{v1}^{(a')}(t-1)+I_{v2}^{(a')}(t-1)\big)\frac{S_{v2}^{(a)}(t-1)}{N_a}-\theta E_{v2}^{(a)}(t-1)
$$

$$
I_u^{(a)}(t) = I_u^{(a)}(t-1)+\theta E_u^{(a)}(t-1)-\gamma I_u^{(a)}(t-1)+n_{imp,u}^{(a)}(t)
$$

$$
I_{v1}^{(a)}(t) = I_{v1}^{(a)}(t-1)+\theta E_{v1}^{(a)}(t-1)-\gamma I_{v1}^{(a)}(t-1)+n_{imp,v1}^{(a)}(t)
$$

$$
I_{v2}^{(a)}(t) = I_{v2}^{(a)}(t-1)+\theta E_{v2}^{(a)}(t-1)-\gamma I_{v2}^{(a)}(t-1)+n_{imp,v2}^{(a)}(t)
$$

$$
R_u^{(a)}(t) = R_u^{(a)}(t-1)+\gamma I_u^{(a)}(t-1)-n_{vacc1}^{(a)}(t-1)\frac{R_u^{(a)}(t-1)}{S_u^{(a)}(t-1)+R_u^{(a)}(t-1)}
$$

$$
R_{v1}^{(a)}(t) = R_{v1}^{(a)}(t-1)+\gamma I_{v1}^{(a)}(t-1)+n_{vacc1}^{(a)}(t-1)\frac{R_u^{(a)}(t-1)}{S_u^{(a)}(t-1)+R_u^{(a)}(t-1)}-n_{vacc2}^{(a)}(t-1)\frac{R_{v1}^{(a)}(t-1)}{S_{v1}^{(a)}(t-1)+R_{v1}^{(a)}(t-1)}
$$

$$
R_{v2}^{(a)}(t) = R_{v2}^{(a)}(t-1)+\gamma I_{v2}^{(a)}(t-1)+n_{vacc2}^{(a)}(t-1)\frac{R_{v1}^{(a)}(t-1)}{S_{v1}^{(a)}(t-1)+R_{v1}^{(a)}(t-1)}.
$$

### 2 Data sources and model parameters

#### 2.1 Reported cases

Data on reported cases and vaccination is collected from the Public Health Agency of Sweden.[1] During 2021 there were 882,000 reported cases, 4,100 admissions to the ICU and 5,540 deaths. The time series of reported cases is seen in Figure S1. The figure shows a strong weekly pattern with lower number of reported cases on weekends. The number of imported cases are relatively larger during July, August when the number of reported cases are low.

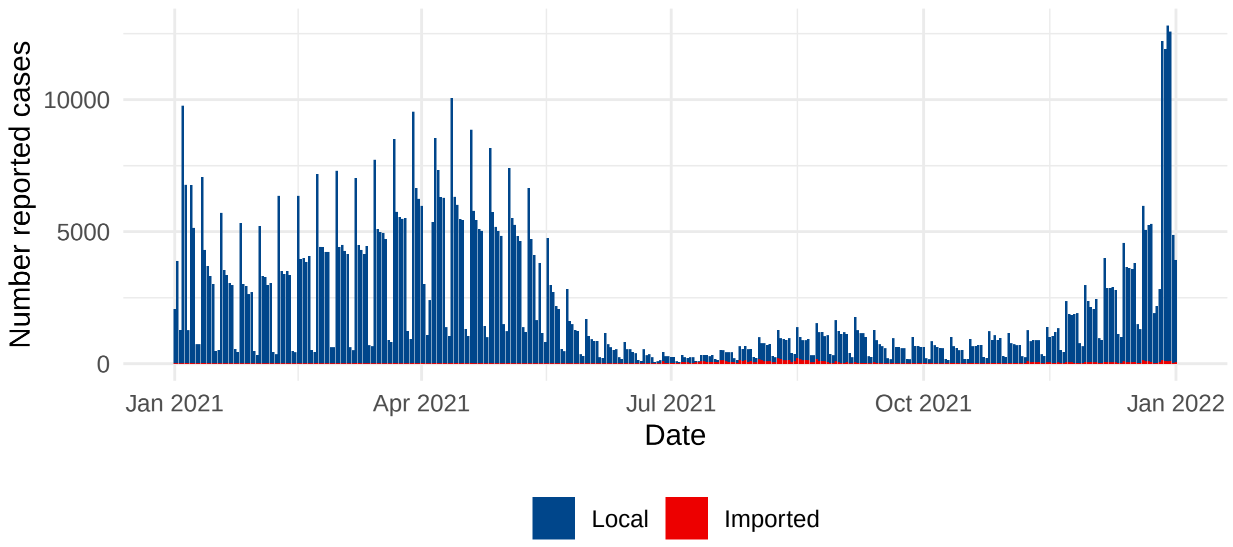


Figure S1: Time series of daily reported local and imported COVID-19 cases for Sweden during 2021.

#### 2.2 Underreporting

The under-reporting of cases depend on the behavior of the population and testing strategy which may vary over time. To account for this in our analysis we use the estimated reporting fraction estimated by the Public Health Agency of Sweden.[2] The estimation includes three age groups; 0-19, 20-69 and 70+ years of age. In Figure S2 the estimates are shown (red) together with splines (green) interpolating between the estimated fractions. There are two different estimates; model free and model based. We will use the model free estimates in the main analysis and the model based in a sensitivity analysis. The estimates of the fraction reported cases are overall lover in the model based estimates. What implication using the model free estimates compared to the model based has for the analysis can be seen in the sensitivity analysis in Section 5.

#### 2.3 IFR

We use age-specific IFR estimates derived from literature.[3−5] We use the combined information from the three sources as no source provide estimates for each age group. The IFR estimates used in our analysis are found in Table S1.

Table S1: IFR (%) used in our analysis.

| Age group | IFR (%) |
|---|---:|
| 20-29 | 0.002 |
| 30-39 | 0.010 |
| 40-49 | 0.040 |
| 50-59 | 0.200 |
| 60-69 | 0.500 |
| 70-79 | 2.00 |
| 80-89 | 7.00 |
| 90+ | 15.0 |

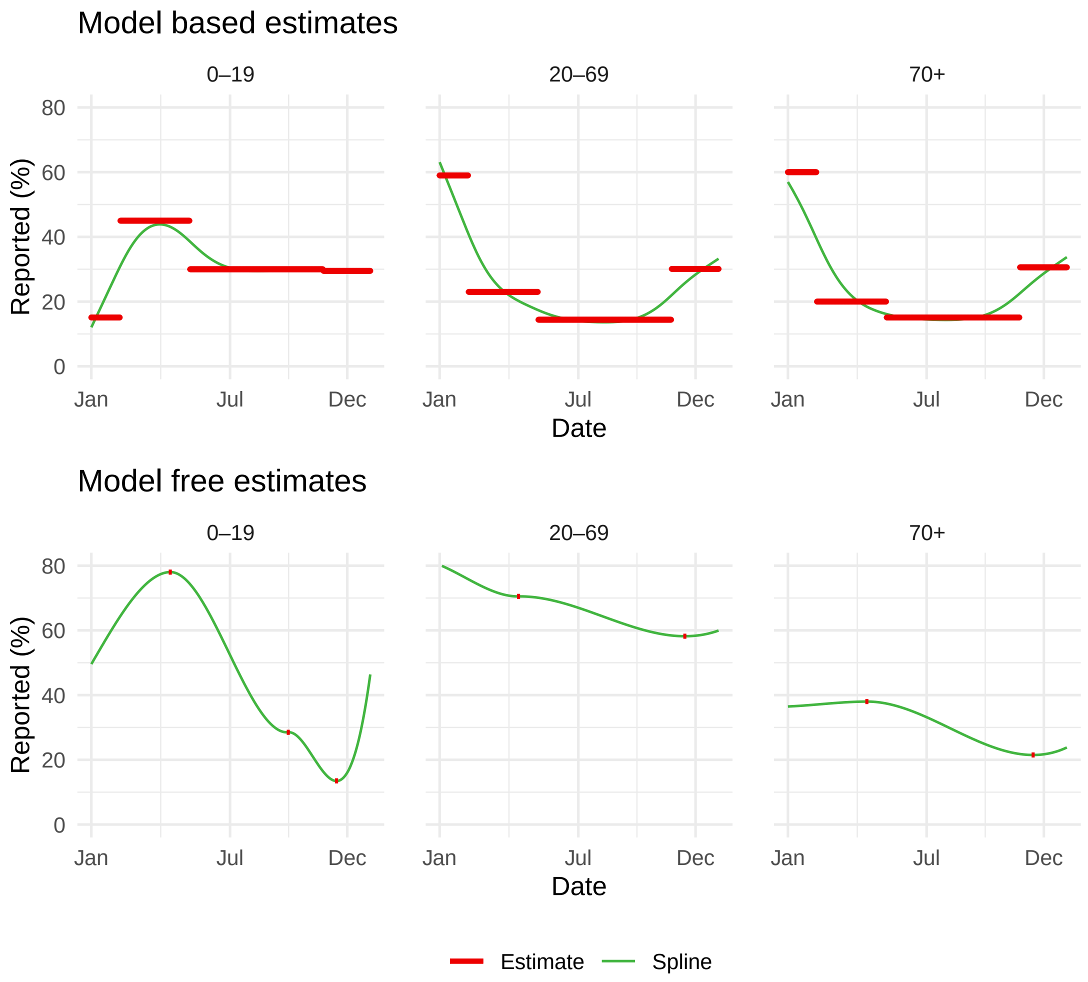


Figure S2: Estimated fraction of reported cases to the number of infections for three different age groups; 0-19, 20-69 and 70+ years of age in Sweden 2021. Red lines are estmations from the Public Health Agency of Sweden and the green lines are splines interpolating between the piecewise constant estimates.

#### 2.4 Parameter values

Parameter values used in our analysis is found in the table below.

Table S2: Parameter values used in our analysis.

| **Parameter** | **Notation** | **Value** |
|---|---|---|
| Incubation period | $\theta^{-1}$ | 4.6 days |
| Duration of infection | $\gamma^{-1}$ | 2.1 days |
| VE infectivity | $ve_{inf}$ | 50% |
| VE susceptability 1 dose | $ve_{susc,v1}$ | 50% |
| VE susceptability 2 doses | $ve_{susc,v2}$ | 80% |
| VE mortality | $ve_{mort}$ | 91% |

#### 3 Statistical analysis

We assume that the number new infections of day $t$ follow a Poisson distribution given by

$$\text{new infections}_{t,a} \sim \text{Pois}(\text{reported cases}_{t,a} * \text{fraction observed}_{t,a})$$

where subscript $\{t, a\}$ denotes day $t$ and age group $a$.

We use a random walk prior for the infectivity rate $\beta(t)$ and priors found in Table S3.

Table S3: Priors for the Bayesian hierarchical model.

| Parameter | Notation | Prior |
|---|---|---|
| Standard deviation of $\beta(t)$ | $\sigma_\beta$ | N(1, 0.2) |
| Log of $\beta(1)$ | $\log(\beta(1))$ | N(0.66, 0.3) |

We run the MCMC chains for 400 iterations using 4 cores.

#### 4 Age-specific results

Figure S3 is complementary to Figure 4A in the main manuscript showing the age aggregated results. We can see that the age stratified reported deaths also follow fairly well our estimated case fatalities from the factual analysis.

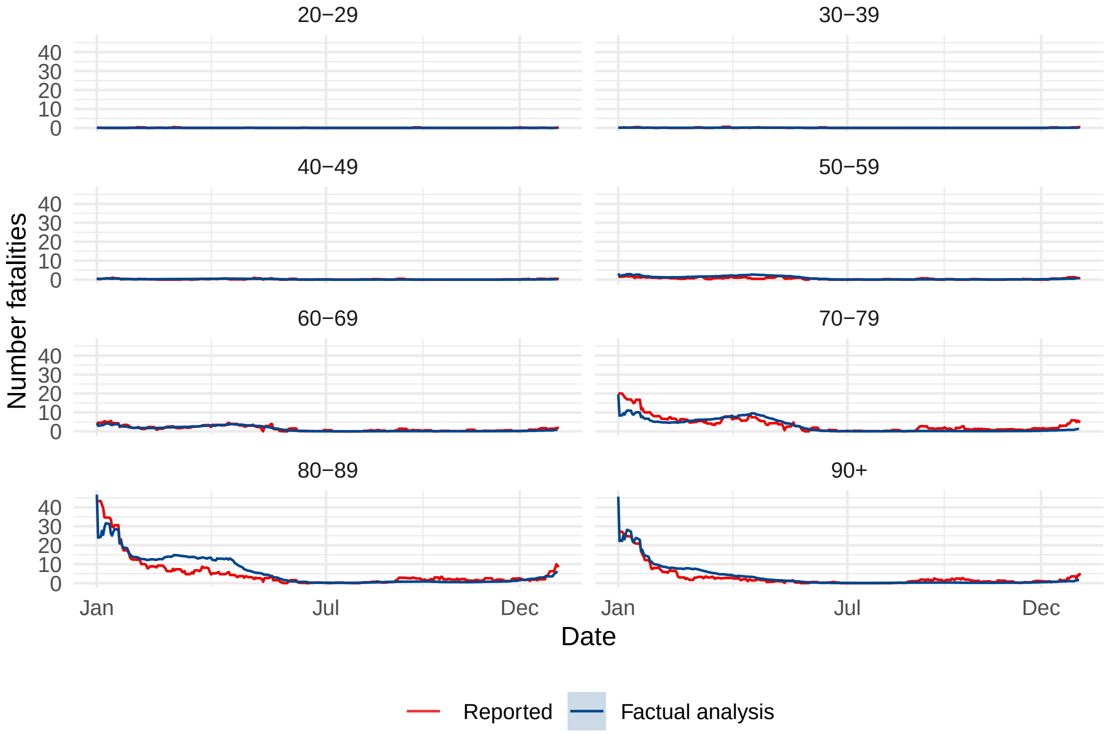


Figure S3: Age stratified reported and estimated case fatalities from factual analysis over time.

#### 5 Details of sensitivity analysis

The model based estimates of the underreporting shifts a large weight of the number of lives saved to the direct effect (as seen in Table 3). The results in terms of estimated number of case fatalities over time can be seen in Figure S4.

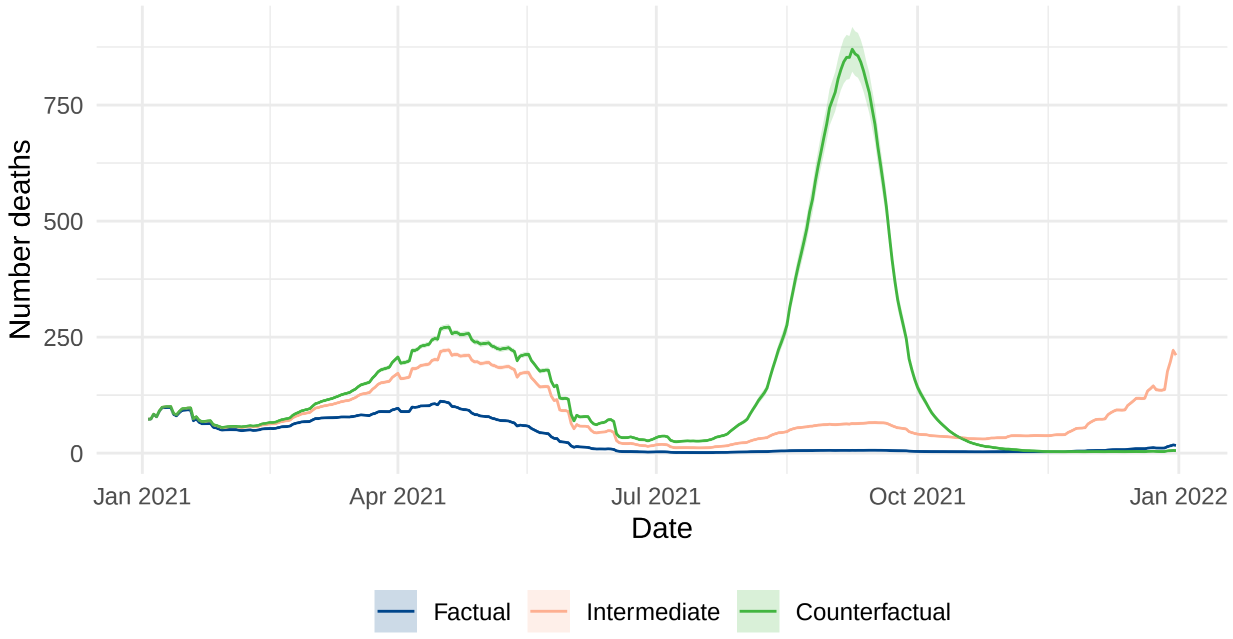


Figure S4: Estimated number of case fatalities from factual, intermediate and counterfactual models using the models based estimates of the under-reporting.

We see a larger number of estimated deaths in spring and smaller in fall compared to our main analysis (seen in Figure 4B in the main manuscript). The estimates of the fraction unreported cases are overall higher in the model based estimates. This implies that it would have been a higher number of infections which would also lead to a higher number of case fatalities. The shift in the effect from indirect to direct effect is mainly driven by a larger fraction of unreported cases in the first part of 2021, when it is mainly the elderly population being unvaccinated.

[2] Public Health Agency of Sweden. 2022. Estimating the fraction unreported covid-19 cases. Available upon request by the public health agency.

[3] Public Health Agency of Sweden. 2020. The Infection Fatality Rate of COVID-19 in Stockholm – Technical Report. https://www.folkhalsomyndigheten.se/contentassets/da0321b738ee4f0686d758e069e18caa/skattning-letalitet-covid-19-stockholms-lan.pdf/.

[4] Pezzullo AM, Axfors C, Contopoulos-Ioannidis DG, Apostolatos A, Ioannidis JPA. 2023. Age-Stratified Infection Fatality Rate of COVID-19 in the Non-Elderly Population. *Environmental Research, 216(Pt 3)*, no. 114655. https://doi.org/20210308.

[5] Rickards CG, Kilpatrick AM. 2023. Age-Specific SARS-CoV-2 Infection Fatality Rates Derived from Serological Data Vary with Income and Income Inequality. *PloS One* 18 (5): e0285612. https://doi.org/10.1371/journal.pone.0285612
